# Functional Impact of Macular Atrophy and Fibrosis in Neovascular Age-Related Macular Degeneration by Defect-Mapping Microperimetry

**DOI:** 10.64898/2026.09.27.26364134

**Authors:** Emily K. Glover, Robyn H. Guymer, Carla J. Abbott, Maxime Jannaud, Xavier Hadoux, Zhichao Wu

## Abstract

**Purpose:** To understand the functional impact of macular atrophy (MA) and fibrosis in eyes with treated neovascular age-related macular degeneration (nAMD) based on deep visual sensitivity losses.

**Design:** Cross-sectional study.

**Participants:** One-hundred and nine eyes from 97 individuals with nAMD that had been treated for ≥12 months without significant retinal fluid.

**Methods:** All participants underwent defect-mapping microperimetry (DMP) testing – a strategy optimized to quantify the spatial extent of deep visual sensitivity losses through single 10 decibel stimuli presentations – where 208 locations were sampled in the central 8º radius region.

Participants also underwent color fundus photography, fundus autofluorescence and OCT imaging, which were co-registered and manually annotated for MA and fibrosis to determine their two-dimensional pointwise extent at individual test locations, and global extent in the corresponding region where DMP was performed.

**Main Outcome Measures:** Pointwise association between the presence of MA and fibrosis with missing a stimulus on DMP and global association between the extent of MA and fibrosis with the proportion of locations missed (PLM) on DMP.

**Results:** At a pointwise level, the presence of MA and fibrosis were independently associated with an increased likelihood of missing a stimulus on DMP (odds ratio = 42.5 and 2.5 respectively; *P* <0.001 for both). However, at a global level, only an increasing extent of MA was associated with a significantly higher PLM on DMP (*P* < 0.001), but not fibrosis (*P* ≥ 0.227). MA extent explained a substantial proportion of the variance in the PLM (*R*^*2*^ = 0.97) and there was a very strong correlation between the two parameters (ρ = 0.93).

**Conclusions:** This study confirmed the expected functional impact of MA and fibrosis on DMP testing at a pointwise level. However, it showed at the global level that only the extent of MA, and not fibrosis, was independently associated with functional loss on DMP, and that there was a very strong structure-function correlation based on MA. These findings thus suggest that evaluating MA alone may be sufficient for capturing structural changes strongly associated with the global extent of deep visual sensitivity losses in those with treated nAMD.

## INTRODUCTION

Following the first introduction of anti-vascular endothelial growth factor (VEGF) treatments for exudative neovascular age-related macular degeneration (nAMD) around two decades ago, significantly fewer people were severely visually impaired compared to the era before such treatments became available.^1^ However, despite anti-VEGF treatment, initial vision improvement or vision stabilization is often not maintained in the longer term. Only approximately half of the eyes with nAMD retain driving-level vision within 5 years of treatment,^2,3^ and only approximately 15% to 35% of eyes retain this level of vision after 10 years.^4-7^

A previous study evaluating a large real-world registry of treatment-naïve nAMD eyes that underwent anti-VEGF treatment showed the development of subfoveal macular atrophy (MA) or subretinal fibrosis was associated with significantly worse visual acuity outcomes over a 36-month period.^8^ These findings are supported by numerous previous studies that have observed that the presence or development of MA and fibrosis are the main factors associated with poor visual outcomes in nAMD.^8-13^

However, previous studies have only observed moderate correlations between VA and the extent of MA and fibrosis,^14,15^ which may reflect the limitations of VA to comprehensively and sensitively capture the functional impact of these pathological changes. To facilitate the development of new treatments to prevent the onset or growth of MA and fibrosis in eyes being treated for nAMD, there is a need to more comprehensively understand the functional impact of these pathological changes in the setting of nAMD.

Microperimetry (or fundus-tracked perimetry) could enable such functional impact to be more effectively characterized. Several previous studies have shown that visual sensitivities measured by conventional threshold-based microperimetry are significantly reduced in nAMD eyes with MA and fibrosis.^16-19^ However, none of these studies have reported the strength of the correlation between visual sensitivity losses and these pathological changes.

Furthermore, we have recently shown that a novel defect-mapping microperimetry (DMP) testing strategy^20^ – optimized to quantify the spatial extent of deep visual sensitivity defects – captured functional losses that showed a very strong correlation with the extent of geographic atrophy (GA).^21,22^ This study thus sought to examine the functional impact of MA and fibrosis with DMP testing, based on current consensus-based definitions of these features,^23,24^ in eyes with nAMD that have been treated for at least 12 months.

## METHODS

This study included individuals enrolled in an observational study of the functional loss in eyes with treated nAMD conducted at the Centre for Eye Research Australia (CERA). This study was conducted in adherence with the tenets of the Declaration of Helsinki and International Conference on Harmonization Guidelines for Good Clinical Practice. Institutional review board approval was obtained for this study, and all participants provided informed consent prior to enrolment.

### Participants

This study included individuals who were ≥50 years old with exudative nAMD in at least one eye, who had undergone anti-VEGF treatment for ≥12 months and with a best-corrected visual acuity (BCVA) of 20/240 or better on the Early Treatment Diabetic Retinopathy Study (ETDRS) letter chart. Eyes with significant retinal fluid (≥10% of the region tested on microperimetry, described further below) were excluded. The other exclusion criteria in this study included the presence of any ocular, systemic, or neurologic condition(s) that could affect the reliable evaluation of visual function or the retina.

### Defect-Mapping Microperimetry

DMP testing was performed using the Macular Integrity Assessment (MAIA) microperimeter (CenterVue, Padova, Italy), as described previously in detail.^21,25-28^ Briefly, testing was performed following pupillary dilation and before any study assessments that could bleach the retina (e.g., CFP or FAF) or compromise the ocular surface. Testing was performed using Goldmann Size III stimuli with an intensity of 10 dB (relative to the dynamic range of the device; stimulus luminance = 32.9 cd/m^2^ and background luminance = 1.27 cd/m^2^), as it corresponds to the floor of the effective dynamic range of the device and thus captures deep visual sensitivity losses characteristic of truly non-responding locations.^29,30^ For DMP testing, such 10 dB stimuli are only presented once at each location, unlike the multiple times a stimulus is presented at each location with standard threshold-based testing. This enables testing to be performed at a much higher spatial density with DMP testing than standard threshold-based testing over a similar test duration.

This study used an isotropic stimulus pattern that sampled the central 8º radius region with 208 test locations and at an inter-stimulus interval of 1º. If fixation was non-foveal, the stimulus pattern was manually centered on the fovea as estimated from the OCT volume scan. This study included participants who completed two reliable DMP tests in a single session, with a reliable test being defined by the presence of ≤25% false-positive errors (derived from catch trials of 10 dB stimuli presented at the optic nerve head).

### Multimodal Retinal Imaging

Non-stereoscopic CFPs centered on the macula with a 45º diameter field-of-view were acquired using the CR6-45NM device (Canon; Tokyo, Japan). Short-wavelength (blue) FAF images of the central 30º × 30º region (with at least 768 × 768 pixels and 100 frames averaged) and OCT volume scans of the central 20º × 20º region (with 97 B-scans, each with 1024 A-scans and 16 frames averaged) were obtained using the Spectralis HRA+OCT device (Heidelberg Engineering GmbH; Heidelberg, Germany).

### Image Annotations and Processing

From the two DMP tests performed per eye, the near-infrared fundus image of the test with the better image quality was selected and, along with the CFPs and FAF images, co-registered with the near-infrared reflectance (NIR) image of the OCT volume scan. This was achieved by manual placement of fiducial points at retinal vessel bifurcations, followed by automated affine (including scaling, rotation, and translation) and radial transformations. The co-registered CFP, FAF, and OCT images were then annotated for MA, fibrosis and retinal fluid – masked to the DMP test results – using a custom software (Cross-Modality Annotation Software [XMAS]; Ophthalmic Neuroscience Unit, Centre for Eye Research Australia).^31,32^

Annotations for MA were performed using a similar process to that used in the analysis of previous clinical trials.^33,34^ Candidate MA lesions were first identified based on regions of definite decreased autofluorescence on the FAF images, and OCT B-scans were then reviewed to confirm the presence of complete retinal pigment epithelium (RPE) and outer retinal atrophy (cRORA) as per a previous consensus-based definition.^23^ Once confirmed, the MA lesion was then annotated on the FAF image, and was required to be ≥175 µm in diameter. Annotations for fibrosis were performed using a two-step process and definitions as per the recently published expert group consensus paper.^24^ Candidate regions of fibrosis were first identified based on regions with well-defined, highly hyperreflective material (HRM) with a laminated or banded appearance that showed associated evidence of RPE disruption on OCT B-scans. CFPs were then reviewed to confirm the presence of a well-defined yellow, white or gray lesion corresponding to the HRM. Once confirmed, the region of fibrosis was then annotated on the CFP, defined by the extent of the HRM present on OCT imaging. The annotations for MA and fibrosis were performed by one grader (E.K.G.) after their presence at the eye level was first determined together with three other investigators of the study (C.J.A., R.H.G., and Z.W.). Annotations for retinal fluid – both subretinal and intraretinal fluid – detected on OCT B-scans were also performed on the NIR image of the OCT volume scan by one grader (E.K.G.). Figure 1 shows examples of eyes annotated for MA and fibrosis. Following image annotations, the global extent of MA, fibrosis and retinal fluid within the 8º radius region sampled on DMP testing, and at each of its 208 test locations, were extracted.

**Figure 1:**
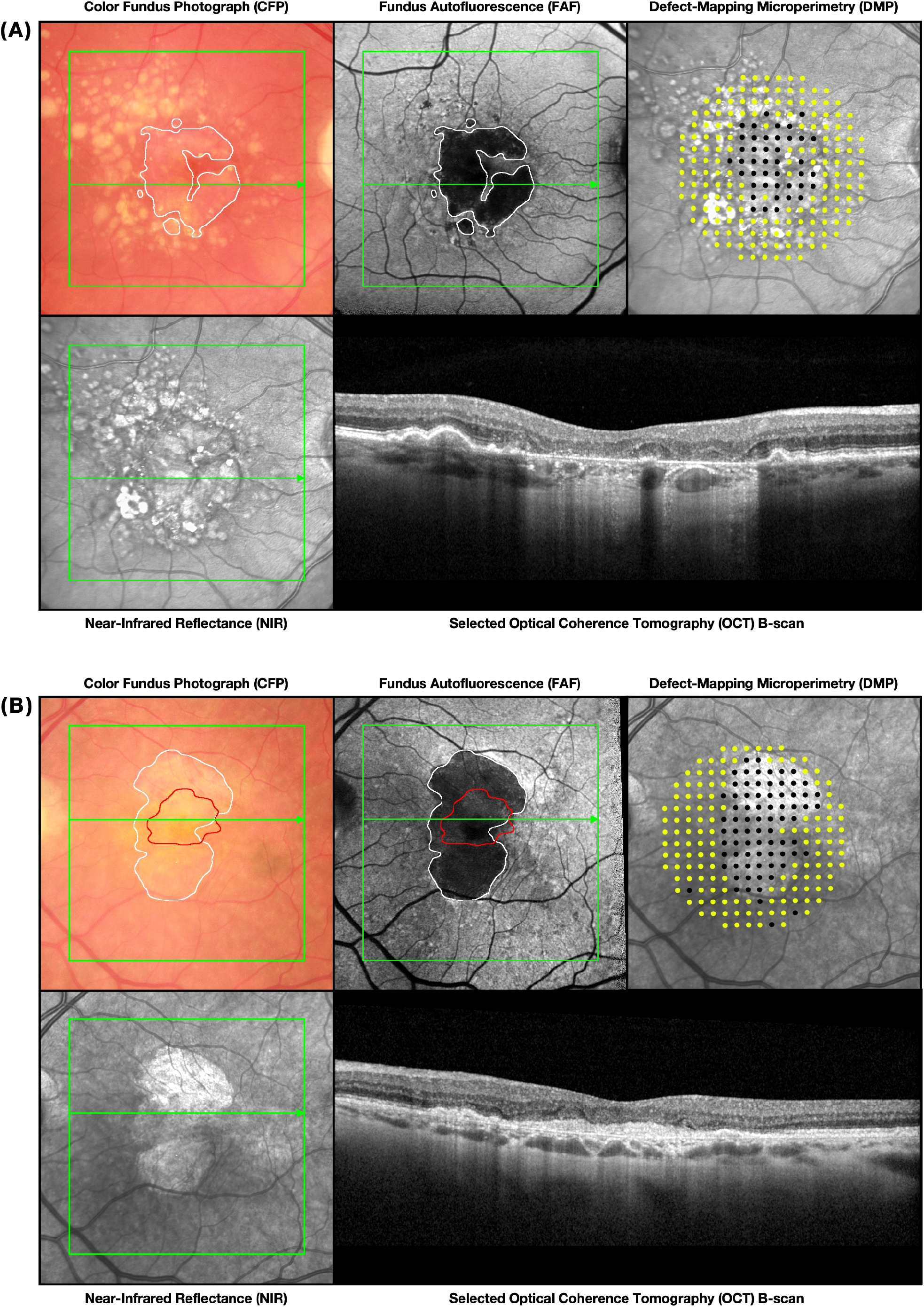
Examples of eyes with (**A**) macular atrophy (MA), and (**B**) both MA and fibrosis, included in this study, showing their color fundus photographs (CFPs), fundus autofluorescence (FAF), defect-mapping microperimetry (DMP) results (208 test locations in the central 8º radius region; yellow markers = seen, black markers = not seen), near-infrared reflectance (NIR) image, and a selected OCT B-scan. The location where the OCT volume scan was taken is indicated by the green squares, and the location where the selected OCT B-scan was taken through is indicated by the green arrows, on the CFP, FAF and NIR images. The area(s) annotated for MA and fibrosis are shown as white and red outlines respectively on the CFP and FAF images.

### Statistical Analyses

The presence of systematic changes in the proportion of locations missed (PLM; from all 208 locations) on DMP testing was evaluated using a random intercepts linear regression model, to account for the correlations between two tests per eye and two eyes per individual. Test-retest repeatability of DMP testing was evaluated based on the intrasession coefficient of repeatability (CoR) of the PLM, representing where 95% of its test-retest differences are expected to lie.

Pointwise structure-function associations were then performed to evaluate if the presence of MA, fibrosis and retinal fluid at a test location (based on feature occupying the entire region sampled by the stimulus) was associated with a significantly higher probability of missing a stimulus on DMP testing, when compared to their absence (in the entire region sampled by the stimulus). This was evaluated using a multivariable mixed-effects logistic regression model, to account for the correlations above and those between multiple locations per test. The probabilities of missing a stimulus based on the presence of MA and fibrosis were also derived to illustrate the magnitude of the associations.

Global structure-function associations were then performed to similarly evaluate if an increasing global extent of MA, fibrosis and retinal fluid in the central 8º radius region was associated with an increasing PLM on DMP testing, using a multivariable random intercepts model to account for the between- and within-eye correlations. The proportion of variance explained (*R*^*2*^) in the PLM by the global extent of MA, and the Spearman rank correlation coefficient (ρ) between these two measures, were also derived and their 95% confidence intervals determined using non-parametric bootstrap resampling at the person level (n = 1,000 bootstrap resamples). All analyses were conducted using Stata software version 18 (StataCorp, College Station, TX).

## RESULTS

This study included 109 eyes from 97 participants, who were on average 80 ± 6 years old (range, 56 to 92 years old) and 74 (76%) participants were female. The eyes included had a median BCVA of 74 letters (interquartile range [IQR] = 61 to 81 letters) read on the ETDRS chart. There were 57 (52%) eyes that had some retinal fluid in the region where DMP testing was performed, and the median *en face* percentage and area of this region occupied by retinal fluid for these eyes was 0.2% (IQR = 0.1 to 0.6%) and 0.03 mm^2^ (IQR = 0.02 to 0.10 mm^2^). There were 86 (79%) eyes with MA and 58 (53%) eyes with fibrosis in the region where DMP testing was performed, and there were 99 (91%) eyes with either MA or fibrosis, and 45 (41%) eyes with both MA and fibrosis.

### Characteristics of Defect-Mapping Microperimetry Testing

There were 81 (74%), 71 (65%) and 63 (58%) eyes that had ≥1, ≥3 and ≥5 repeatably non-responding test locations respectively. The median duration of the DMP tests was 5.9 minutes (IQR = 5.6 to 6.4 minutes) when considering the entire cohort, and 6.1 minutes (IQR = 5.8 to 6.5 minutes) when considering only eyes with ≥3 repeatably non-responding test locations. There was no significant systematic difference in the PLM between the first and second test when considering the entire cohort (*P* = 0.989), nor when considering only eyes with ≥3 repeatably non-responding test locations (*P* = 0.472). The intrasession CoR of the PLM was 5.3% (95% CI = 4.3 to 6.3%) when considering the entire cohort, and 6.1% (95% CI = 4.7 to 7.5%) when considering only eyes with ≥3 repeatably non-responding test locations.

### Pointwise Structure-Function Association

At a pointwise level, the presence of MA, fibrosis and retinal fluid at a tested location was associated with a significantly higher probability of missing a stimulus on DMP (*P* ≤ 0.004 for all; Table 1). To illustrate the magnitude of these associations, the pointwise probability of missing a stimulus based on the presence of MA and fibrosis are presented in Table 2.

**Table 1.** Multivariable analysis of the pointwise likelihood of missing a stimulus on defect-mapping microperimetry.

| Presence of Feature | Odds Ratio | P-Value |
| --- | --- | --- |
| Macular atrophy | 42.5 (38.5 to 46.9) | < 0.001 |
| Fibrosis | 2.5 (2.1 to 2.8) | < 0.001 |
| Retinal fluid | 6.5 (1.8 to 23.5) | 0.004 |

**Notes:** The presence of a feature is defined when it occupies the entire region sampled by the stimulus, and it is compared to the absence of the feature in this entire region. Values in the parentheses represent the 95% confidence intervals.

**Table 2:** Pointwise probability of missing stimuli (%) on defect-mapping microperimetry testing based on the presence or absence of fibrosis, when macular atrophy was either absent or present, at the location sampled.

| Macular Atrophy | Fibrosis |  | Odds Ratio | P-Value |
| --- | --- | --- | --- | --- |
|  | Absent | Present |  |  |
| Absent | 8 (6 to 10) | 17 (13 to 20) | 2.8 (2.3 to 3.5) | < 0.001 |
| Present | 63 (58 to 68) | 73 (68 to 78) | 1.8 (1.5 to 2.3) | < 0.001 |

**Notes:** The presence of a feature is defined when it occupies the entire region sampled by the stimulus, and it is compared to the absence of the feature in this entire region. Analyses were performed adjusting for the presence of retinal fluid at the tested location. Values in the parentheses represent the 95% confidence intervals.

### Global Structure-Function Association

At a global level however, an increasing extent of MA was associated with a significantly higher PLM on DMP (*P* < 0.001), but not an increasing extent of fibrosis or retinal fluid (*P* ≥ 0.227) in a multivariable analysis; these findings are summarized in Table 3.

**Table 3:** Multivariable analysis of the global association between the proportion of locations missed (%) on defect-mapping microperimetry (DMP) and extent of macular atrophy (MA), fibrosis and retinal fluid.

| Parameter | Coefficient | P-Value |
| --- | --- | --- |
| MA extent (per 10% increase) | 9.6 (9.3 to 9.9) | < 0.001 |
| Fibrosis extent (per 10% increase) | 0.3 (-0.3 to 0.8) | 0.355 |
| Fluid extent (per 10% increase) | -0.8 (-8.6 to 7.0) | 0.839 |

**Notes:** All measurements of extent are considered within the 8° radius region sampled by DMP. Values in the parentheses represent the 95% confidence intervals.

A scatterplot of the PLM against the extent of MA is shown in Figure 2, where MA explained a substantial proportion of the variance in the PLM (*R*^*2*^ = 0.97 [95% CI = 0.95 to 0.98]) and there was a very strong correlation between the two measures (ρ = 0.93 [95% CI = 0.88 to 0.96]).

**Figure 2:**
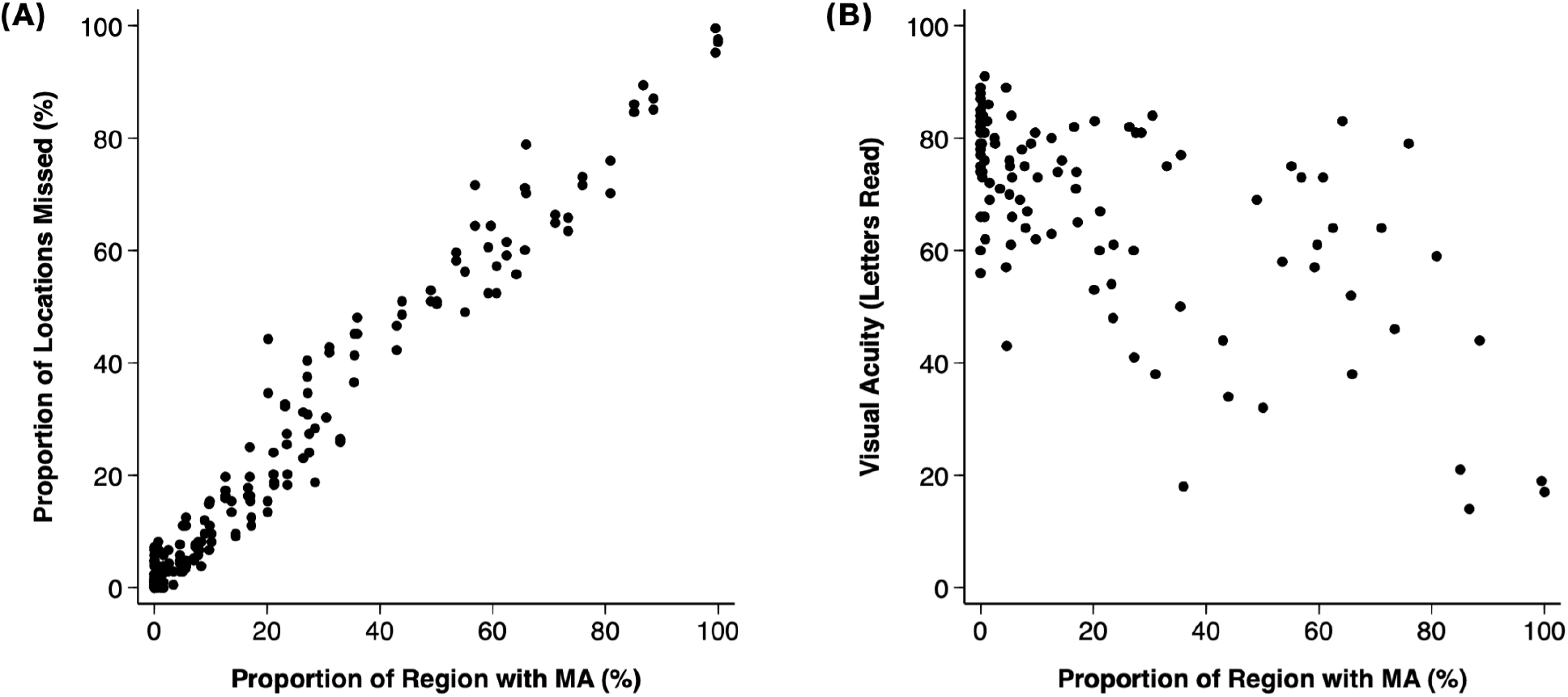
Plot of (**A**) the proportion of locations missed on defect-mapping microperimetry (out of the 208 locations tested), and (**B**) best-corrected visual acuity (number of letters read on the on the Early Treatment Diabetic Retinopathy Study [ETDRS] chart), against the proportion of the region sampled with macular atrophy (MA).

When considering BCVA for reference, an increasing extent of MA was also associated with a significantly reduced BCVA (*P* < 0.001), but not an increasing extent of fibrosis and retinal fluid (*P* ≥ 0.156) in a multivariable analysis. MA explained a lower proportion of the variance in the BCVA (*R*^*2*^ = 0.41 [95% CI = 0.23 to 0.58]) and showed a weaker correlation with BCVA (ρ = -0.59 [95% CI = -0.72 to -0.45]) than seen with the PLM. A scatterplot of BCVA against MA extent is also shown in Figure 2.

## DISCUSSION

This study showed that at a pointwise level, the presence of MA and fibrosis were both associated with a significantly higher probability of missing a stimulus on DMP. At a global level however, only the extent of MA and not fibrosis, remained independently significantly associated with the PLM, and MA was very strongly correlated with this functional outcome. These findings thus confirm the expected functional impact of these two features, but they also demonstrate how evaluation of MA alone may be sufficient for capturing the global extent of functionally relevant structural changes.

The findings of this study that the presence of MA and fibrosis were independently associated with an increased probability of missing a stimulus on DMP at a pointwise level are broadly consistent with those from previous studies reporting a significant reduction in visual sensitivities on threshold-based microperimetry in association with the presence of MA and fibrosis.^16-19^ Roh and colleagues^16^ reported that both atrophy (defined on OCT) and fibrosis were independently associated with reduced visual sensitivities, but their study included only 15 eyes with late AMD (including both late neovascular and atrophic AMD) and did not provide a definition of fibrosis. Schranz and colleagues^17^ evaluated 30 eyes of 30 consecutive individuals with nAMD treated for ≥1 year and observed that the presence of fibrosis as detected on different imaging modalities were associated with worse visual sensitivities. However, the coexisting impact of MA was not considered in this study, and the proportion of eyes with fibrosis were also not reported. Bygglin and colleagues^18^ reported that the presence of FAF-defined MA was associated with reduced visual sensitivities in a study of 24 eyes of 24 individuals with nAMD, but they did not examine fibrosis. Finally, Tan and colleagues^19^ reported that OCT-defined atrophy and fibrosis were both independently associated with reduced visual sensitivities in a cohort of 228 eyes from 120 individuals, of whom 106 eyes had nAMD. However, they performed a multivariable logistic regression analysis of mean visual sensitivity despite it being a continuous outcome measure, and thus the validity of their findings cannot be determined.

Note that none of these previous studies reported the strength of the structure-function correlations to provide insights into how strongly MA and fibrosis account for the functional impact in eyes with nAMD. This study instead showed for the first time that only the extent of MA, and not fibrosis, was significantly and independently associated with the global PLM on DMP testing. This study also showed that MA explained 97% of the variance in this functional outcome and that a very strong structure-function correlation exists between these two parameters (ρ = 0.93). These findings are largely consistent with our previous observations with DMP testing in eyes with GA, where we found that the extent of FAF-defined GA and OCT-defined RPE, ellipsoid zone (EZ) and external limiting membrane (ELM) loss were also all strongly correlated with the PLM on DMP testing (ρ = 0.85 to 0.89).^21,22^

The findings that the extent of fibrosis was no longer independently associated with the PLM on DMP are accounted for by the observations that the presence of fibrosis was associated with statistically significant increased probability of missing a stimulus at a pointwise level, but that the magnitude of this increase was dramatically smaller compared to MA (odds ratio = 2.5 and 42.5 respectively). It is also worth noting that in this study, 46% of DMP test locations with fibrosis present in the entire region sampled by a stimulus had some degree of MA in that region sampled. As such, it is possible that the functional loss seen in association with fibrosis in previous studies, that considered fibrosis alone without evaluating MA, could be associated with coexistent MA rather than the independent effect of fibrosis without MA.^15,17^

The findings of this study thus suggest that evaluation of MA alone may be sufficient when seeking to capture structural changes that are highly correlated with the global extent of deep visual sensitivity losses. The findings of this study should not be taken to suggest that fibrosis is not functionally relevant, but rather that at a global level, deep visual sensitivity losses seen in association with fibrosis are largely accounted for by coexistent MA. In addition, whilst the presence of fibrosis can complicate the quantification of MA,^35^ the findings of this study suggest that the evaluation of MA with an approach similar to that used in the analysis of previous clinical trials^33,34^ enables quantification of structural changes that are closely associated with functional losses. Together, these findings provide crucial insights into the functional impact of MA and fibrosis – as defined using consensus-based definitions of these features^23,24^ – that can support future work seeking to develop new treatments to prevent the onset or growth of these pathological changes.

Limitations of this study include the evaluation of only nAMD eyes that have been treated for ≥12 months and without significant retinal fluid, which was specifically performed to understand the functional impact of MA and fibrosis in relatively stable and inactive nAMD, and to minimize the confounding impact of exudative disease activity in nAMD. As such, the generalizability of the findings of this study to eyes that have been treated for a shorter duration or with more extensive exudation is not known. Note also that this cohort does not represent a consecutive series of individuals meeting the eligibility criteria in this study, as individuals with nAMD were referred for this prospective observational study with the stated goal to understand the functional impact of MA and fibrosis. As such, there was a referral bias for individuals with such features, as reflected by the high prevalence (>90%) for either feature in this cohort. Another limitation of this study is that the image annotations were only performed by one grader. Although not performed for every eye included in this study, these image annotations were also reviewed by the other three investigators of this study for most of the eyes, especially when there was uncertainty about the presence and extent of MA and fibrosis. Annotations were then revised as required following a group discussion. Future studies are thus needed, ideally involving multiple readers from different reading centers, to determine the reproducibility of these findings. Further work is also needed to examine if the findings would differ based on the grading process and definitions used for MA and fibrosis, such as if MA or fibrosis was quantified using FAF and CFP alone respectively, or if fibrosis was defined in a less stringent manner than the recent consensus definition.^24^ Further work is also needed to examine if the very strong structure-function correlation seen with manually annotated MA would similarly be seen with automatically segmented OCT outer retinal band loss.^22^

In conclusion, this study confirmed the expected functional impact of MA and fibrosis based on DMP testing by showing that their presence is associated with a significantly higher probability of missing a stimulus at a pointwise level. However, it showed that only the extent of MA, but not fibrosis, was independently associated with the global extent of functional loss on DMP, and that MA extent was very strongly associated with this functional outcome. These findings thus suggest that the evaluation of MA alone may be sufficient for capturing structural changes that are strongly associated with the global extent of the deep visual sensitivity losses present in eyes with treated nAMD.

## Data Availability

Data not available for this manuscript.

